# “Cost effectiveness analysis and effects of treatment on the years of life with disability among the adult cleft lip and cleft palate patients in Bangladesh”

**DOI:** 10.1101/2025.08.01.25332786

**Authors:** Tanzila Rafique, Shahriar Mohd Shams, Md. Zahid Hasan

## Abstract

Oro facial clefts pose a large psychological and economic burden for the families and society. The number of years lost in a person’s life caused by these disabilities can be assessed by calculating the Years lived with disability (YLD), a component of the Disability Adjusted Life Years (DALY) method. It is used for the evaluation of economic impact and to quantify the disease burden. Over a period of four years, this cross-sectional study was carried out at the orthodontics Department of BSMMU, Bangladesh. 55 adult male and female patients of 15-30 years were enrolled in the study. Everyone completed their treatment at the time of the interview. The number of DALYs averted after the treatment of these patients was calculated. Cost effectiveness of treatment was assessed by comparing the cost per DALY averted with the WHO-provided GDP threshold. If the patients were untreated, they would have a total of 1,314.35 years lived with disability, considering the disease onset at 0 years of life up to the expected standard life expectancy of 72 years. Because of the patients’ treatment, a total of 372.41 years of living with disability (YLD) have been averted. Our study found that the available treatments for clefts are cost-effective in our country. As the economic gain that calculated, focused on a small proportion of the DALY averted and cost-effectiveness, it advocates for broader implementation of such a method to address other health interventions.

## Introduction

Orofacial clefts (OFC) are the most common birth malformations. About one in five to seven hundred newborns are born with OFC^1^. The potential for lifelong morbidity, its complex etiology, and the substantial multidisciplinary effort needed for intervention make cleft lip and palate a serious public health concern. The socioeconomic well-being, quality of life, and health of affected individuals and their families may be profoundly impacted by certain birth anomalies. Additionally, they lead to significant healthcare costs and utilization.

In Indian and Oriental ethnicities, these craniofacial abnormalities are more common, affecting about 1 in 700 infants ^2^. Oro-facial clefts were identified as one of the primary anomalies in a community-based birth defects study carried out in rural Bangladesh^3^.

Cleft-born babies in Bangladesh have an annual economic impact of USD 250 million on the country’s GDP ^4^. A multidisciplinary team of surgeons, speech therapists, audiologists, dentists, orthodontists, psychologists, geneticists, and specialized nurses treat cleft lip/cleft palate in developed countries. Primary surgery to close the defect, continued orthodontic and speech therapy, and additional and tertiary surgeries to improve the initial surgical outcome are all components of the optimal treatment plan. The first six months of life are typically when the major surgical treatment is carried out.

There is a serious lack of skilled surgeons and anesthesiologists in developing countries who are capable of conducting surgery to deal with this medical condition. Therefore, there is often no preoperative or postoperative therapy. Out of the 120 million people living in Bangladesh, about 300,000 have cleft lip and palate (CLAP), and they frequently come from low-income families ^5^. The great majority of these patients do not have access to and are unable to pay for even the most basic surgical treatments or services related to clefts. According to statistics, the predicted life span of an individual with an untreated cleft is 14 years shorter than the national average. Managing a person who was born with a cleft requires the assistance of several professions ^6^.

Bangladesh lacks population health summary measures that aggregate data on non-fatal health outcomes to demonstrate a population’s overall health in one statistic. A variety of indicators have been developed over the last thirty years or so, that adjust mortality in order to compensate for the impact of illness or disability. Many countries are currently looking into the potential utilization of DALYs as a tool for cost-effectiveness studies, priority setting, and as a measure of trends in disease burden ^6,7^.

Like in high-income countries (HICs), policymakers in low- and middle-income countries (LMICs) face challenging decisions about the most effective way to utilize the health care resources at their disposal for the purpose of accomplishing established social targets. Cost-effectiveness analysis—more particularly a system according to the GDP of a country per capita has been approved by WHO to aid in policymaking ^8^. Regional differences in cost, demography, and resource use are demonstrated by the wide range in cost-effectiveness for similar procedures.

Health interventions seek to avert DALYs, and in doing so, raise the number of years that a person lives in good health. This study assessed the effects of cleft treatment on years of life with disability among the adult Cleft lip/palate patients by estimation of the number of DALYs averted after the treatment of the selected patients in our country. This also assessed the financial viability of available treatments of Cleft lip/palate by comparing with the WHO-provided GDP threshold.

## Materials and methods

### Study design and setting

This cross-sectional study was conducted from the department of Orthodontics, Faculty of Dentistry of Bangabandhu Sheikh Medical University (BSMMU), Shahbag, Dhaka, Bangladesh from 07/11/2010 to 25/10/2024, over four years. Sample was collected purposively from the BSMMU, Shahbag, Dhaka, Bangladesh; Sheikh Hasina National Institute of Burn and Plastic Surgery (SHNIBPS), 63, AHM Kamruzzaman Sharani, Chankharpul, Dhaka, Bangladesh, and Bangladesh Specialized Hospital, 21 Shyamoli, Mirpur Road, Dhaka-1207, Bangladesh. The World Health Organization Study on Global Ageing and Adult Health (SAGE) provided the questionnaire required for obtaining the socio-demographic information ^9^. By comparing the cost per DALY averted with the GDP threshold set by the WHO, the cost-effectiveness of the treatment was evaluated.

### Participants

The study sample consisted of 55 adult treated male and female patients aged 15 to 30. The participants were chosen based on the inclusion and exclusion criteria. To finalize the research tools and materials, five participants were chosen for the pre-testing.

### Key variables ascertainment

Before commencement, the study along its objective was explained to all participants.

### Orofacial clefts

An opening or split in the upper lip that results from an unborn baby’s developing facial tissues unable to close fully completely is called a cleft lip. An opening in the roof of the mouth, known as a cleft palate, develops when tissue fails to fuse during fetal development ^10^.

### DALY

A disability-adjusted life year, or DALY, is a metric used to quantify health burden that includes both decreased life expectancy and a lower quality of life. The equation DALY = YLL + YLD is the mathematical representation of a DALY. One DALY is equivalent to the loss of one year of good health. The human capital approach, which calculates DALYs, makes the assumption that individuals are like machines and that lost years of life are equal to lost years of productivity.

### YLL (Years of life lost)

Years of life lost, or YLL, is a metric used to quantify the reduction in life expectancy.

### YLD (Years lived with disability)

Years lived with disability, or YLD, is a metric that measures the diminished quality of life which an individual with an illness or injury experiences.

### Cost per DALY averted

Cost-effectiveness analyses that measure and exhibit the costs and effects of programs and at least one alternative in a ratio of incremental cost to incremental effect are referred to as cost-per-disability-adjusted life year (DALY) averted (or cost-per-DALY) studies ^12^.

### Disability weight

Years lived with disability (YLD) for specific health outcomes in a population are calculated using disability weights, which represent the extent of health loss associated to these outcomes. A scale from 0 to 1 is used to measure the weights, with 0 denoting perfect health and 1 denoting death are used to calculate years lost to disability (YLD) 13.

### GDP (Gross domestic product)

This measures the monetary value of final goods and services—that is, those that are bought by the final user—produced in a country in a given period of time ^14^.

### VSL

It measures the extent that society values the lowering of mortality. The value of the small reduction in mortality risks can be simply summarized using the VSL. It is not intended to be used to the value of converting a person’s risk of death from one to zero, or saving their life ^15^.

### GNI (Gross National Income)

It measures the overall income generated by individuals and companies in a nation, inclusive of investment income, no matter the location of its generation. Additionally, it includes funds received from outside the country, such as foreign investments and economic assistance. This serves as an indicator of national wealth that can act as a substitute for gross domestic product (GDP). To compute GNI, incorporate income from international sources into a nation’s GDP ^16^.

### Ethical considerations

The ethical approval was obtained from the IRB of Bangabandhu Sheikh Medical University (BSMMU) before the commencement of the study. Informed written consent and assent were obtained from the guardians and participant, explaining the purposes of the study, goals, minimal risks, and benefits. A separate identification number was given to each patient to maintain confidentiality. Participation in the study was voluntary, and participants had the complete right to withdraw themselves at any time for the study without hampering their regular treatment procedures.

### Statistical analysis

Data were analyzed by IBM SPSS Statistics for Windows, Version 20.0. (Armonk, NY: IBM Corp). Descriptive statistics in the form of frequency distribution and percentage were used to explain the background characteristics of the study participants. The YLD metric of DALY (DCPI-the Disease Control Priority Project) ^17^ was employed to calculate the total number of years lost as a consequence of disability. YLD was calculated by multiplying the number of new cases of a disease (I) by the disability weight (DW) with the average amount of time a person had suffered from it before it goes away or is remitted. The disability weight for cleft lip/palate was obtained from the life table of disease burden used in the modified DCPI in order to measure the DALYs and to reflect the lifelong burden of disease. To determine how many DALYs were averted due to the patients’ therapy, we employed the WHO-provided DALY calculation method. In order to evaluate the number of years left to live with a reduced burden of disease, the age at treatment was compared with WHO life expectancy data, and each patient’s age and sex were included at surgery^18^. By comparing the cost per DALY averted with the GDP threshold given by the WHO, the economic effectiveness of the current therapies for each patient was evaluated. A treatment is considered very cost-effective if the cost per DALY averted is lower than one-tenth of GDP per capita, according to the WHO-provided GDP threshold ^19^.

## Results

### Background characteristics of patients

This study included 55 adult patients with cleft lip/palate who had completed their treatment by the time of their interviews.

**Error! Reference source not found**. provides frequency (n) distribution and percentage (%) of patients by age, gender, marital status, education, occupation, income source, monthly household income, and living area types. The highest proportion of patients (52.7%) was in the 25–29 age category, followed by the 15–24 age group (26%), and the 30+ age group (21.8%). The patient group consisted of 38.2% females and 61.8% males, with an average age of 27 years. Of the patients, 60.0% were unmarried, 29.1% were married, and 10.9% were widowed or divorced.

In the treated group, 31% of patients were unemployed, 35% were students, 78.2% relied on family or other income sources, and 21.8% had personal income. Sixty-nine percent of the families of patients had a monthly income exceeding BDT 15,000. Ten percent of the patients resided in rural locations, twenty-four percent in peri-urban settings, and sixteen percent in urban environments. (TABLE 1).

**TABLE 1:** Background characteristics of study participants.

| Characteristics | Treated |  |
| --- | --- | --- |
| *Age group* | n | % |
| 15 to 24 years | 14 | 25.5 |
| 25 to 29 years | 29 | 52.7 |
| 30 and above | 12 | 21.8 |
| **Mean age in years (SE)** | 55 | 27.25 (0.57) |
| *Gender* |  |  |
| Female | 21 | 38.2 |
| Male | 34 | 61.8 |
| *Marital Status* |  |  |
| Unmarried | 33 | 60.0 |
| Married | 16 | 29.1 |
| Others (divorced, widow) | 6 | 10.9 |
| *Education* |  |  |
| Up to the primary education | 1 | 1.8 |
| Secondary incomplete | 6 | 10.9 |
| Secondary school completed | 6 | 10.9 |
| University/College completed | 24 | 43.6 |
| Postgraduate | 18 | 32.7 |
| *Occupation<sup>1</sup>* |  |  |
| Student | 19 | 34.5 |
| Employed | 19 | 34.5 |
| Unemployed | 8 | 30.9 |
| **Income source<sup>1</sup>** |  |  |
| Own income | 12 | 21.8 |
| Family/others | 43 | 78.2 |
| **Monthly Family income** |  |  |
| Less than BDT 10000 | 4 | 7.3 |
| BDT 10000 to BDT 15000 | 4 | 7.3 |
| more than BDT 15000 | 38 | 69.1 |
| No fixed income/ uncertain | 9 | 16.4 |
| **Living area** |  |  |
| Urban | 36 | 65.5 |
| Peri urban | 13 | 23.6 |
| Rural | 6 | 10.9 |

### Effect of cleft treatment on years of life with disability among patients

#### DALYs averted from treatment among cleft lip/palate patients

TABLE 2 presents the estimation of the amount of DALYs averted following the treatment of the chosen patients. The 55 patients who were enrolled in this study sought healthcare for issues related to cleft lip/palate, and at the time of the interview, they had completed their treatment. We employed disability weights to determine the DALYs for individuals with cleft palate (0.231 without treatment and 0.15 with treatment) and cleft lip (0.098 without treatment and 0.016 with treatment). To calculate the number of DALYs averted due to the patients’ treatment, we utilized the DALY calculation method provided by the WHO. Given that the illness began when the patients were born and the average life expectancy is 72 years, we estimated that, without treatment, all patients would have experienced impairment over a cumulative total of 1,314.35 years.

**TABLE 2.** Number of DALYs averted after treatment of cleft lip and cleft palate patients.

| Type of patients by age | Standard life expectancy at this group | Duration of illness | Number of patients | DALYs without treatment | DALYs after treatment | DALY averted |
| --- | --- | --- | --- | --- | --- | --- |
| **Male** |  |  |  |  |  |  |
| 20-24 | 57.9 | 26 | 13 | 358.90 | 236.18 | 122.72 |
| 25-29 | 53.0 | 27 | 16 | 421.13 | 282.90 | 138.23 |
| 30-34 | 48.0 | 31.5 | 4 | 104.58 | 73.31 | 31.28 |
| 35-39 | 43.1 | 38.5 | 1 | 26.84 | 19.82 | 7.02 |
| **Subtotal** |  |  | **34** | **911.5** | **612.2** | **299.2** |
| **Female** |  |  |  | - | - | - |
| 20-24 | 60.5 | 26 | 1 | 19.99 | 16.05 | 3.93 |
| 25-29 | 55.7 | 27 | 13 | 248.47 | 201.37 | 47.10 |
| 30-34 | 50.7 | 31.5 | 4 | 75.94 | 62.76 | 13.18 |
| 35-39 | 45.9 | 38.5 | 3 | 58.50 | 49.55 | 8.95 |
| **Subtotal** |  |  | **21** | **402.9** | **329.7** | **73.2** |
| **Total** |  |  | **55** | **1,314.35** | **941.94** | **372.41** |

However, the total number of years lived with a disability would decrease to 941.94 once therapy had been finished at a different age. Therefore, a total of 372.41 years of living with disability (YLD) have been averted because of the treatment of the cleft patients.

#### Cost-effectiveness of cleft lip/palate treatment

Based on TABLE 3, the average cost for treating a patient with a cleft lip/palate was BDT 420,827. This comprised direct medical expenses like surgery (BDT 363,000), medication (BDT 43,833), and follow-up care (BDT 3,760). The total costs also consisted of direct non-medical costs, which were the transportation costs for the patients and/or caregivers of BDT 10, 234 per case also made up the overall expenses. According to this calculation, the overall cost for treating all patients amounted to BDT 23,145,500. According to Table 1, the treatment of cleft patients resulted in a total of 372.41 DALYs being averted. Consequently, the cost per DALY averted was calculated to be BDT 62,150, equivalent to US$730.55 for the treatment of both cleft lip/palate patients, which is below one times the GDP per capita (US$2458).

**TABLE 3.** Cost per patient for cleft lip and cleft palate patients and cost per DALYs averted.

| Components | Amount in BDT |
| --- | --- |
| **Direct medical expenditure** |  |
| Surgery | 363000 |
| Medicine | 43833 |
| Follow up | 3760 |
| **Direct non-medical expenditure** |  |
| Transport | 10,234 |
| **Total** | 420,827 |
| Total costs of treatment for 55 patients | 23,145,500.62 |
| Total DALYs averted | 372.41 |
| Cost per DALY averted | 62,150 |
| In US\$<sup>a)</sup> | 730.55 |

<sup>a)</sup> Exchange rate of 2021 US\$ = 85.07 BDT

## Discussion

Cost-utility or cost-benefit analysis constitutes cost-effectiveness evaluations. Cost-utility analysis is an economic evaluation approach that involves an overall health measure, such as Disability-Adjusted Life Years (DALYs) ^20^, is employed to value health outcomes. The findings are presented as a dollar-per-DALY-averted incremental cost-effectiveness ratio. Health outcomes are measured in monetary terms in cost-benefit evaluations, and their value can be based on each nation’s Gross National Index per capita (GNI/capita) ^21^. The outcomes are shown as a net benefit or loss or as a cost-benefit ratio.

Assessing the cost per disability-adjusted life years (DALYs) retained with treatments is a method that is becoming more common to evaluate the cost-effectiveness of community health improvement initiatives.

These studies have surged substantially in recent years, with many of them concentrating on health interventions in countries with low or middle incomes.

In our study, we calculated the cost per DALY averted for 55 adult patients treated for cleft lip and palate. The disability severity weights utilized in this research range from 0 (indicating no negative impact on quality of life) to 1 (representing a burden comparable to the preference for death), and are applied to adjust Years Lived with Disability (YLD) ^23^.

This study revealed that the overall number of years lived with a disability will reduce to 941.94 following treatment completion at different ages. Because of the cleft patients’ treatment, 372.41 years of living with disability (YLD) have been averted.

In the 2001 report^24^ on Macroeconomics and Health, Jeff Sachs proposed that a cautious evaluation of the economic impact of disease could be achieved by measuring Disability Adjusted Life Years (DALYs) in relation to Gross National Income per capita, a method often referred to as the human capital approach.

Utilizing patient data gathered from an Interplast site in Nepal, Corlew subsequently utilized this approach to calculate the economic benefit of each individual resulting from CLP repair ^25^. Although he attributed an economic benefit to CLP repair using the VSL methodology, he did not give it the same credence as the human capital approach.

A study in sub-Saharan Africa, where a large portion of the population lacks general access to good surgical care, integrated the two approaches, the value of a Statistical Life (VSL) approach and the human capital approach. The number of DALYs that would be averted if each case were surgically corrected at birth was taken into account to calculate the economic value of cleft corrections. Disability weights (DW) were obtained from the 2004 global burden of disease study^26^. This study found the DW were 0.098 for cleft lip and 0.231 for cleft palate; treatment reduces these to 0.016 and 0.015, respectively^27^.

In our research, the cost per Disability-Adjusted Life Year (DALY) averted for the treatment of patients with cleft lip and palate was found to be US$730.55, which is below the GDP per capita of US$2458 (ref). According to the World Health Organization’s GDP criterion, a treatment is regarded as highly cost-effective if its cost per DALY averted is under one times the GDP per capita. Consequently, this treatment was rated as cost-efficient.

Another study that measured the burden of disease prevented by the worldwide surgical efforts of a major cleft charity discovered the cost-effectiveness of treatment. Over a ten-year span, they calculated the economic impact of this endeavor. Disability-adjusted life years (DALYs) and anonymized data from all primary cleft lip and cleft palate operations in the Smile Train database were examined and computed in this study. In addition to the country-specific life expectancy tables, predefined disability weights, estimated surgical success, and residual disability probability, they also included other age weighting and discounting permutations. Averted DALYs were calculated, and gross national income (GNI) per capita was then multiplied by averted DALYs to estimate economic gains ^28^.

The Quality-adjusted life-years (QALY) are another measure of years lived in perfect health gained. This method lacks sensitivity and may be difficult to apply to chronic disease and preventative treatment ^29^. Both the QALY and DALY methods incorporate the severity and duration of illness or injury and the age at which it occurs. QALY and DALY-based ratios for the same intervention can differ; differences tend to be modest and are unlikely to materially affect resource allocation recommendations. On the other hand, the modest differences may still affect the decision-making process when considered from a broader perspective, including the opportunity cost of other healthcare interventions, budgets for healthcare spending, and price negotiation ^30^.

While both QALYs and DALYs can evaluate cost-effectiveness to aid in healthcare decision-making, additional research is required to implement these measures in addressing specific local health needs and issues. Both QALY and DALY measures cannot fully capture the wider effects from interventions, such as the emotional and mental health, impact on careers and family, non-health effects such as economic and social consequences (e.g., loss of work). In our study, we collected the costs related information retrospectively from the participants. Thus, there might be some chance of recall bias in the collected information. It was the first study that assessed the cost-effectiveness of cleft treatments by using the DALY approach in our country. The quantification of the burden of cleft lips and palates within the DALYs framework found in this study was significant, and it will allow the treatment of clefts to be compared with other health interventions. It can also be used for evidence-based health care resource allocation. The cost-effectiveness analysis could therefore be an important measure for the policy makers to consider when deciding how to fund the delivery of care.

## Conclusion

The economic evaluation of our study demonstrates that the cleft lip and palate treatments are cost-effective in our country. Therefore, these treatments are need to be made accessible and available all over the country. Though the economic gain calculated in this study focused on a small proportion of the DALY averted and the cost-effectiveness of cleft treatment, it can be further applied to evaluate interventions for other health conditions.

## Acknowledgements

We are grateful to the study participants for their cooperation and active participation, which helped to achieve the desired outcomes. We are grateful to the Department of Orthodontics, Faculty of Dentistry of Bangabandhu Sheikh Medical University (BSMMU), Shahbag, Dhaka, Bangladesh, Sheikh Hasina National Institute of Burn and Plastic Surgery (SHNIBPS), 63, AHM Kamruzzaman Sharani, Chankharpul, Dhaka, Bangladesh, and Bangladesh Specialized Hospital, 21 Shyamoli, Mirpur Road, Dhaka-1207, Bangladesh, for their cooperation to achieve the desired study results.

## Ethical approval

Ethical clearance has been taken from the IRB of the Bangabandhu Sheikh Mujib Medical University (BSMMU), Date: 12-03-2018, No. - BSMMU/2018/3022.

## Data availability statement

We confirm that the data supporting the findings of this study will be shared upon reasonable request.

